# A new enzymatic assay to quantify inorganic pyrophosphate in plasma

**DOI:** 10.1101/2022.05.29.22275726

**Authors:** Stefan Lundkvist, Fatemeh Niaziorimi, Flora Szeri, Matthew Caffet, Sharon F. Terry, Gunnar Johansson, Robert S. Jansen, Koen van de Wetering

**Affiliations:** Department of Dermatology and Cutaneous Biology, Jefferson Institute of Molecular Medicine and PXE International Center of Excellence in Research and Clinical Care, Sidney Kimmel Medical College, Thomas Jefferson University, Philadelphia (PA), USA; Department of Chemistry (BMC), Uppsala University, Uppsala, Sweden; Research Centre for Natural Sciences, Institute of Enzymology, Budapest, Hungary; Department of Biochemistry, Semmelweis University, Budapest, Hungary; PXE International, Damascus (MD), USA; Department of Microbiology, Radboud University, Nijmegen, The Netherlands

## Abstract

Inorganic pyrophosphate (PPi) is a crucial extracellular mineralization regulator. Low plasma PPi concentrations underlie the soft tissue calcification present in several rare hereditary mineralization disorders as well as in more common conditions like chronic kidney disease and diabetes. Even though deregulated plasma PPi homeostasis is known to be linked to multiple human diseases, there is currently no reliable assay for its quantification. We here describe an PPi assay that employs the enzyme ATP sulfurylase to convert PPi into ATP. Generated ATP is subsequently quantified by firefly luciferase-based bioluminescence. An internal ATP standard was used to correct for sample-specific interference by matrix compounds on firefly luciferase activity. The assay was validated and shows excellent precision (<3.5%) and accuracy (93-106%) of PPi spiked into human plasma samples. We found that of several anticoagulants tested only EDTA effectively blocked conversion of ATP into PPi in plasma after blood collection. Moreover, filtration over a 300,000 Da molecular weight cut-off membrane, reduced variability of plasma PPi and removed ATP present in a membrane-enclosed compartment, possibly platelets. Applied to plasma samples of wild type and *Abcc6*^-/-^ rats, an animal model with established low circulating levels of PPi, the new assay showed lower variability than the assay that was previously in routine use in our laboratory.

In conclusion, we here report a new and robust assay to determine PPi concentrations in plasma, which outperforms currently available assays because of its high sensitivity, precision, and accuracy.

## Introduction

Extracellular inorganic pyrophosphate (PPi) is a key regulator of mineralization^1^. Disturbances in extracellular PPi homeostasis underlie several ectopic calcification disorders^2^. A complex system of proteins mediating cellular ATP efflux and ecto-nucleotidases together regulate PPi homeostasis in the extracellular environment, including blood plasma^2,3^. All PPi detected in plasma is derived from ATP released from cells into the extracellular environment^4^. Mechanisms involving the plasma membrane proteins hepatic ATP-binding cassette subfamily C member 6 (ABCC6) and the more ubiquitous Ankylosis homologue (ANKH) account for over 80% of all PPi in plasma^5–8^. ATP released into the extracellular environment is rapidly converted by ecto-nucleoside pyrophosphatase/phosphodiesterase 1 (ENPP1) into adenosine monophosphate (AMP) and PPi^4,6–8^. Tissue Non-specific Alkaline Phosphatase (TNAP) is the major enzyme involved in degradation of PPi to inorganic phosphate (Pi)^9^. An additional layer of regulation is provided by CD73, which converts AMP into adenosine and Pi. Adenosine inhibits expression of TNAP-encoding *ALPL*. Absence of CD73 activity therefore reduces local formation of adenosine, increases TNAP activity and, consequently, results in low PPi plasma levels^10,11^.

Mutations in each of the genes participating in plasma PPi homeostasis have been linked to specific mineralization disorders, as detailed in a recent review from our group^2^. Low plasma PPi levels not only underlie the ectopic calcification seen in rare genetic ectopic mineralization disorders, but also in more common conditions like diabetes and chronic kidney disease^12^. Importantly, arterial calcification is prevalent and an independent risk factor of early death in patients suffering from cardiovascular disease^13,14^.

Despite its crucial role in the prevention of calcification of soft connective tissues and arteries, no assay is currently available to reliably quantify PPi in plasma and other tissues. As a result, vastly different values for normal plasma PPi concentrations in both animal models and humans have been reported^5,6,12,15–21^. In addition, several of the currently available assays are cumbersome, time consuming, involve the use of radionuclides, or are not very robust^12,22–25^.

We here describe the validation of a sensitive bioluminescence-based assay for the reliable quantification of PPi in a wide range of biological matrices, in as little as 5 minutes. This novel assay is highly sensitive, consumes only limited sample, provides reproducible results, and is very robust.

## Materials and Methods

### Animals and preparation of rat plasma

The *Abcc6*^-/-^ (Strain A17) and wild type Sprague Dawley rats were bred in house and received water and food ad libitum^26^. Animals were 8-19 months old at the time of blood sampling. Blood was collected by cardiac puncture using a Vacutainer blood collection needle (product # 368607, BD) in CTAD tubes (product # 367947, BD), containing citrate, theophylline, adenosine and dipyridamole^6^. Directly following blood collection, 50 µL of a 15% K_3_EDTA (w/v, product #: 11991, Alfa Aesar) solution was added. Plasma was prepared by centrifugation (10 min, 4 °C). Collected plasma was filtered over a membrane with a molecular weight cutoff (mwco) of 300,000 Da using Centrisart I devices (Sartorius) as previously described^6^, and stored at −20 °C until analysis. Animal studies were approved by the Institutional Animal Care and Use Committee of Thomas Jefferson University in accordance with the National Institutes of Health Guide for Care and Use of Laboratory Animals under approval number 02135-1.

### Preparation of human plasma

To determine the effect of anticoagulant on plasma PPi concentrations, blood of 3 volunteers was collected in CTAD, EDTA (product # 367841, BD) and heparin (product # 366664, BD) tubes by puncture of the antecubital vein with a Vacutainer Blood Collection Set (product # 367287). In addition, the effect of a combination of CTAD and EDTA was studied by collecting blood in CTAD tubes and adding 50 µL of a 15% K_3_EDTA solution directly following collection. Plasma was prepared after centrifugation for 10 min at 1000 RCF at 4 °C. To assess the effect of filtration on plasma PPi concentrations, part of the plasma was filtered using Centrisart I filtration devices (Sartorius, mwco 300,000 Da) at 2250 RCF for 20 minutes at 4 °C, as done previously^6^. After collection, plasma was stored at −20 °C till analysis. The Institutional Review Board of Genetic alliance gave ethical approval for the studies involving blood collection of human participants (Protocol number JKVDW001).

### Quantification of ATP

A commercially available luciferase/luciferin mix (SL-reagent, Biothema, Sweden) was used to quantify ATP concentrations in plasma. The content of one vial (Prod. No. 11-501-TP) was dissolved in 10 mL of ultrapure water. For each analysis, 100 µL of the dissolved SL reagent and 400 µL TE buffer (2 mM EDTA, 10 mM Tris, pH 7.75, Biothema, Sweden) were mixed in a 3.5 mL round polypropylene cuvette (product # 68.752, Sarstedt, Germany) and introduced into a Sirius tube luminometer (Berthold, Germany).

The ATP concentration was determined by adding 5 µL sample to 500 µL SL-reagent/TE buffer mixture, resulting in an increase in luminescence. Next, 5 µL of a 10 µM ATP standard (Biothema, Sweden) was added to the cuvette and again the increase in luminescence was determined. The ratio of the increase in luminescence (Relative Light Unit, RLU) after sample and standard addition was used to calculate the ATP concentration. All measurements used to quantify ATP were within the linear range of the FB12 Sirius luminometer (< 3,000,000 RLU). When the addition of 5 µL of the 10 µM ATP standard resulted in RLU values > 3,000,000 RLU, the assay was repeated using SL reagent that was further diluted with TE buffer until an RLU value below 3,000,000 was obtained.

### Determination of the combined ATP and PPi concentration

Adenosine 5’-phosphosulfate (APS) and PPi can be converted into ATP by the enzyme ATP sulfurylase. ATP can subsequently be sensitively quantified using the enzyme firefly luciferase, which uses ATP and luciferin to generate a light signal. Of note, in a given sample the ATP formed out of PPi and APS adds to the ATP that was already present in the sample at the start of the reaction. Hence, the combined presence of ATP + PPi is quantified using this approach. The assay mixture to quantify ATP and PPi contained APS (6 µM, Santa Cruz Biotechnology, TX), ATP sulfurylase (ATPS, 150 mU/mL, New England Biolabs, Ma), apyrase (250 mU/mL, “ATP removal reagent”, Biothema, Sweden), and SL-reagent (130 µL/mL, Biothema, Sweden), and was incubated overnight at room temperature to degrade contaminating ATP and PPi. This procedure was used to reduce background and increase sensitivity. Notably, the concentration of apyrase (“ATP degradation reagent”) was very low, hence overnight incubation was needed to remove background PPi and ATP from the reagents. Low apyrase concentration was chosen to not affect ATP detection in the samples. The assay mixture was subsequently stored at −20 °C until use. For each analysis, 500 µL of the assay mixture was put into a 3.5 mL polypropylene cuvette (Sarstedt, Germany) and background luminescence was determined in a Sirius tube luminometer (Berthold, Germany). Next, five (5) µL of sample was added and all PPi in the sample was allowed to be converted into ATP by ATPS as continuously monitored by following the luminescent signal. When all PPi had been converted into ATP and the luminescent signal did not further increase (endpoint), 5 µL of a 10 µM ATP standard (Biothema, Sweden) was added and luminescence was monitored in the Sirius luminometer. The ratio between the increase in luminescence after the addition of the sample and the increase seen after the addition of the ATP standard was used to calculate the combined concentration of ATP and PPi. Finally, the PPi concentration in the sample was calculated by subtracting the ATP concentration from the combined ATP and PPi concentration.

### Assay Validation

The precision of the assay was determined (CV%) with analytical PPi standards of 1.0 µM, 0.1 µM and 0.010 µM (BioThema, Sweden), all in water. Each PPi concentration standard was analysed 6 times (n=6). The accuracy of the assay was evaluated by determining the recovery of the PPi standard added to human plasma. For these analyses a 10 µM PPi standard solution was added to human filtered CTAD/EDTA plasma to increase the concentration by 1.0 µM. A 2.5 µM PPi standard solution was used to increase PPi concentrations in plasma by 0.1 µM. Each spiked plasma sample was analysed 3 times. Stability of PPi and ATP concentrations in plasma prepared from blood using different anticoagulants was determined at room temperature. PPi and ATP concentrations were quantified daily or every other day for up to 9 days. Interday variability was estimated from data obtained in the same experiment in filtered plasma prepared from blood in which CTAD, EDTA or CTAD/EDTA was used as anticoagulant.

The performance of the new assay was compared with the assay used in our previous work ^5,6^ in rat plasma, collected from wild type and *Abcc6*^-/-^ animals^26^.

## Results

Figure 1 delineates the concept of the optimized ATPS-based assay to quantify PPi in biological samples. Addition of the PPi standard resulted in increased luminescence, which reached its maximum after 20-30 seconds. At this point the ATP standard was added, resulting in an immediate increase in luminescence. The PPi concentration was calculated from the increase seen by the addition of PPi relative to that caused by the addition of the ATP standard. Due to the glow-type bioluminescent reagent used in the assay, signals induced by the addition of PPi and ATP were very stable over time.

**Figure 1.**
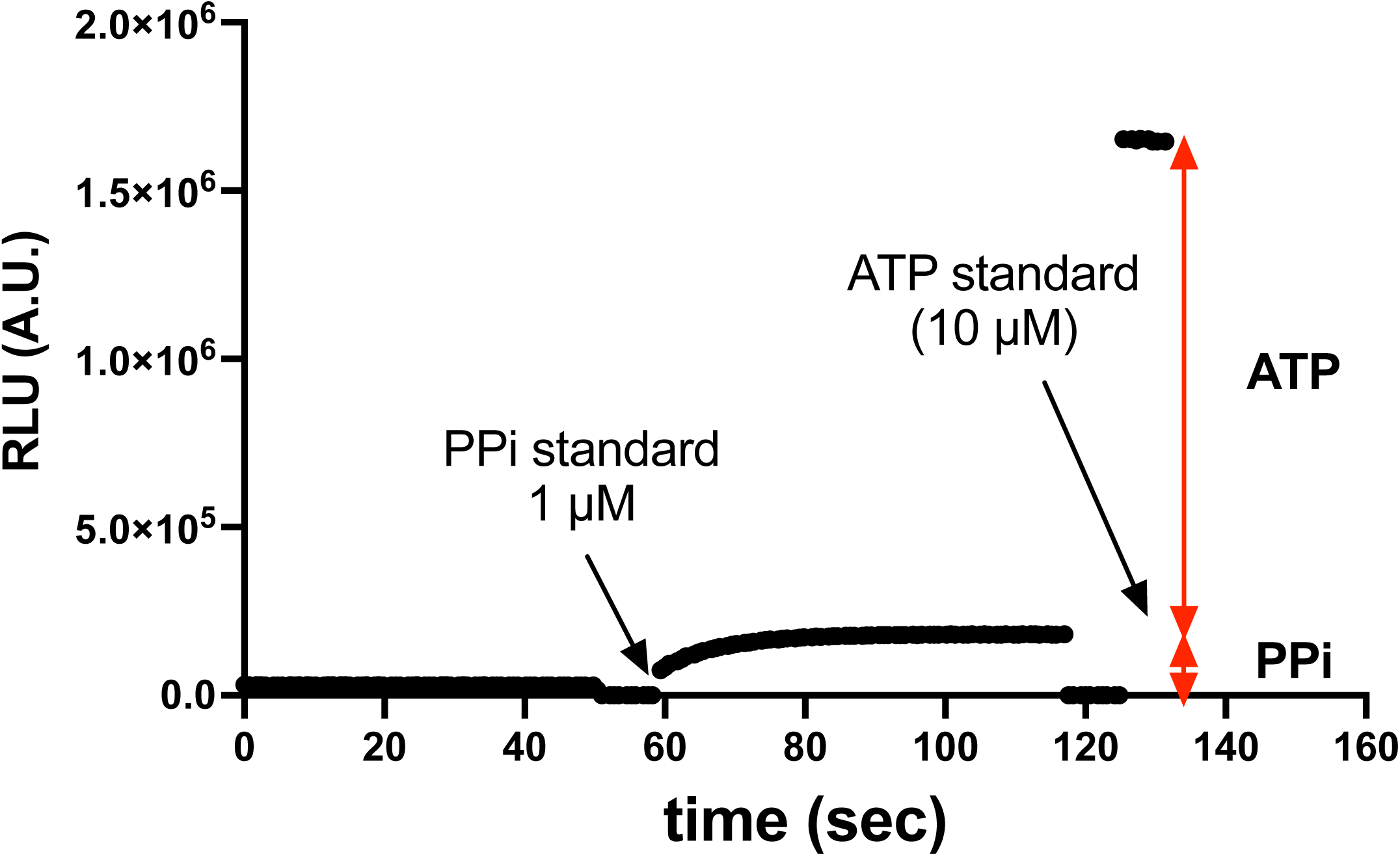
Concept of quantification of PPi using ATP sulfurylase (ATPS). To a reaction mixture containing ATPS, luciferase and luciferin, a 1 µM PPi is added (arrow) and conversion into ATP is allowed to proceed till completion. Next, ATP standard is added and the PPi concentration is calculated from the ratio in increase of luminescence induced by addition of PPi and ATP, respectively. Luminescence is continuously followed in a luminometer. PPi and ATP were added after shortly opening the luminometer. RLU: Relative Light Units.

We first determined the linearity of the FB12 Sirius luminometer with ATP in concentrations ranging from 0.08-10 µM (**Fig. 2A**). Light detection by the FB12 Sirius luminometer was highly linear with a deviation of 3% and 10% at 3,000,000 and 6,000,000 relative light units (RLU), respectively. As expected, the assay was highly linear for concentrations ranging from 0.08 to 10 µM ATP. PPi first needs to be converted into ATP for subsequent detection by luciferin/luciferase. Figure 2B shows that the assay was highly linear for PPi concentrations ranging from 0.08 to 5 µM, indicating efficient conversion by ATPS for the tested concentrations.

**Figure 2.**
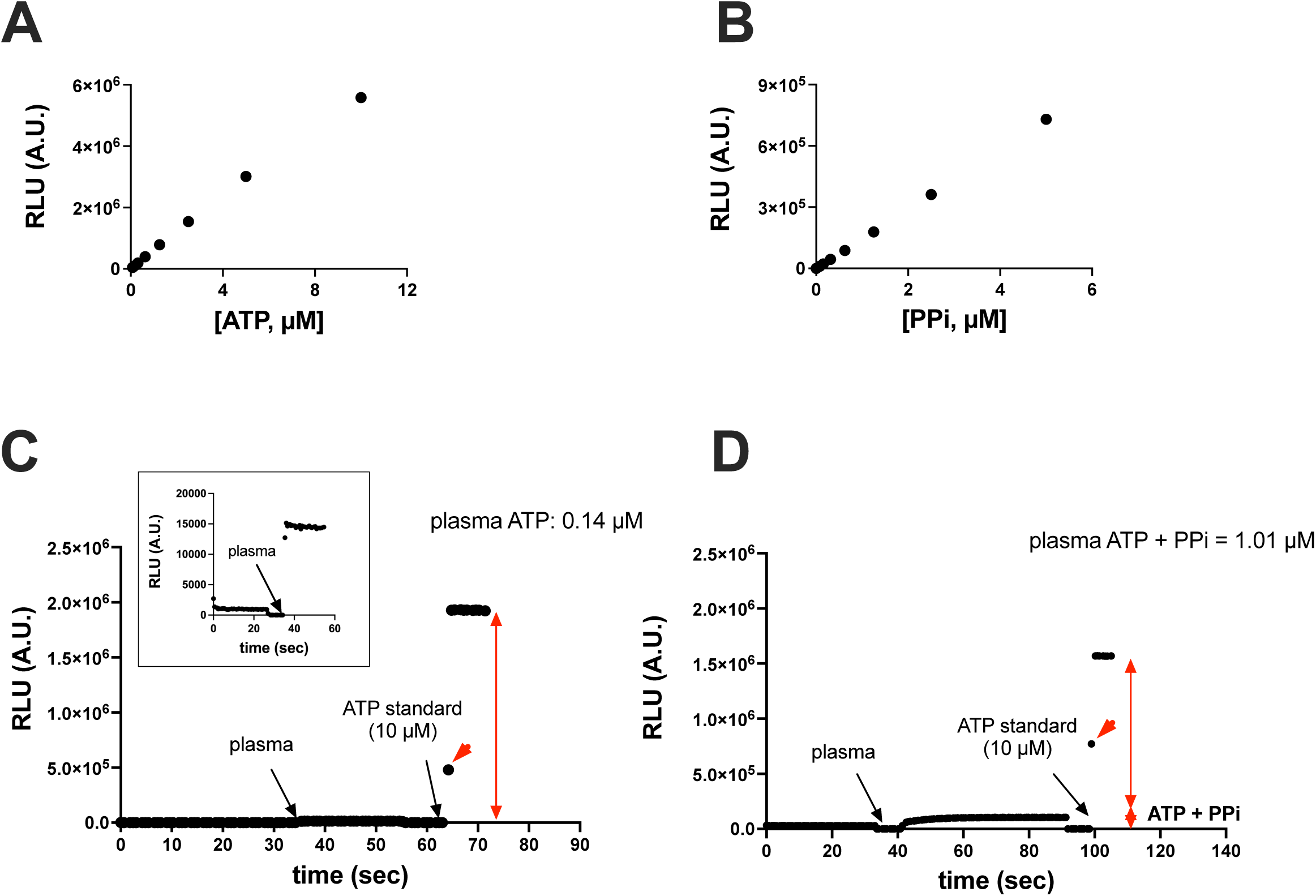
Linearity of luminescence induced by addition PPi and ATP and application to a human plasma sample. Increasing concentrations of ATP (**A**) or PPi (**B**) were added to the reaction mixture, generating signals that were highly linear. In panels **C** and **D**, the concentration of ATP and of ATP + PPi together were determined in a filtered plasma sample containing CTAD and EDTA as anticoagulants. The PPi concentration was calculated by subtracting the ATP concentration (panel C) from the combined ATP and PPi concentration (panel D). Of note, the first data points after addition of ATP (red arrowheads) are lower due to the shortened integration time after closing the luminometer. RLU: Relative Light Units.

To assess the suitability of the assay for plasma PPi quantification, we next tested the assay’s performance on a human plasma sample (filtered CTAD/EDTA). Figure 2C (ATP) and 2D (ATP + PPi) show the time-course of RLU development upon sample addition. The inset in Figure 2C zooms into the increase in luminescence after adding the plasma sample, which would otherwise be masked by the large increase in luminescence seen after addition of the ATP standard. The PPi concentration in the plasma sample shown in Fig 2 was calculated to be 0.87 µM: the combined concentration of PPi and ATP of 1.01 µM (**Fig. 2D**) minus the 0.14 µM ATP detected (**Fig. 2C**).

Next, the precision of the assay was evaluated using 1 µM, 0.1 µM and 0.01 µM PPi standards (Table I). Even at a PPi concentration as low as 0.01 µM, the assays showed good precision with a CV of 8.3%. It is important to note that normal PPi concentrations in plasma range from 1-3 µM, indicating that our assay is able to detect reductions in physiological PPi concentrations accurately.

**Table I:** Precision (SD) and accuracy (deviation) of the PPi assay. Values are calculated from 6 replicate determinations.

|  |  | PPi-standard (μM) |  |  |
| --- | --- | --- | --- | --- |
|  |  | 1.000 | 0.100 | 0.010 |
| Average | μM | 1.011 | 0.102 | 0.010 |
| SD | σ | 0.0141 | 0.0023 | 0.0008 |
| CV | % | 1.40 | 2.31 | 8.32 |
| Accuracy (deviation) | % | 1.09 | 1.70 | 1.56 |

To further evaluate the performance of our assay in biological samples, we determined the recovery of PPi spiked into plasma of three different individuals at 1 and 0.1 µM (Table II). We found an excellent recovery ranging from 97-106% of the added PPi. These analyses demonstrated that the assay was not substantially affected by plasma constituents and provides robust and accurate results. These analyses also confirmed the excellent precision of the assay in biological samples, with CV% ranging from 1.57 to 3.46% for experiments in which 0.1 µM PPi was spiked into the plasma samples. Notably these CV% are not only affected by the precision of the assay but are also influenced by the additional pipetting that is needed to add the PPi standard to the plasma samples.

**Table II:** Recovery of PPi spiked into human plasma indicates excellent accuracy of the PPi assay. Plasma of three individuals was spiked with the indicated amounts of PPi. Afterwards, the total concentration of PPi was determined in the spiked samples. Values are based on triplicate determinations.

|  |  | Individual 1 |  | Individual 2 |  | Individual 3 |  |
| --- | --- | --- | --- | --- | --- | --- | --- |
| **Plasma PPI concentration** | **μM** | 2.163 |  | 1.761 |  | 1.390 |  |
| **PPI added** | **μM** | 1.00 | 0.10 | 1.00 | 1.00 | 1.00 | 0.10 |
| Average PPI concentration detected in spiked plasma | μM | 3.132 | 2.256 | 2.740 | 1.861 | 2.404 | 1.496 |
| SD | σ | 0.0385 | 0.0780 | 0.0768 | 0.0292 | 0.0392 | 0.0385 |
| CV | % | 1.23% | 3.46% | 2.80% | 1.57% | 1.63% | 2.57% |
| PPI recovery (accuracy) | μM | 0.969 | 0.093 | 0.979 | 0.100 | 1.013 | 0.106 |

A soluble form of ENPP1 is present in plasma^27^, which can convert ATP released by, for instance, erythrocytes or platelets into AMP and PPi^3^. Precautions are therefore needed to prevent ENPP1-mediated conversion of ATP into PPi after blood sampling. ENPP1 requires Mg^2+^ for its activity^27^. Mg^2+^ chelating agents, like EDTA and citrate, can therefore be expected to block ENPP1 activity. Platelets are another potential source of PPi released into plasma after blood has been collected. Platelets not only release ATP upon their activation, but also PPi^28^, which might further contribute to artificially high plasma PPi concentrations. Platelet activation probably explains why serum contains much higher PPi concentrations than plasma (not shown). We determined the effects of the type of anticoagulant used and platelet removal on ATP and PPi concentrations in plasma samples of 3 human subjects. In all heparinized plasma samples ATP was undetectable, indicating rapid degradation of ATP by Mg^2+^-dependent ectonucleotidases (**Fig. 3A**). Hardly any ATP was found in filtered plasma, irrespective of the anticoagulant used. This result indicates that the ATP detected in unfiltered plasma is present in a membrane-enclosed compartment, which is removed by filtration. Surprisingly, the platelet-rich plasma did not contain more ATP than unfiltered plasma. For unknown reasons, CTAD and CTAD/EDTA plasma of participant 1 contained relatively high concentrations of ATP. The PPi amount detected depended on the anticoagulant used in unfiltered plasma, and CTAD and heparin yielded higher concentrations. The same was seen for the platelet-rich plasma samples. The type of anticoagulant used did not affect PPi concentrations to the same extent in filtered, platelet-free, plasma samples. All plasma had been stored at −20 C prior to analysis. Possibly, the increased levels of PPi in CTAD and heparin plasma are the result of release of ATP from a membrane-enclosed compartment and its subsequent conversion into PPi, after a freeze-thaw cycle. We did not see the same increase in PPi when EDTA was used as an anticoagulant. These findings indicate that the citrate in CTAD chelates Mg^2+^ less well than EDTA and is unable to block ENPP1-mediated conversion of released ATP into PPi immediately. Heparinized plasma tended to give the highest PPi concentrations, which can be explained by the continued conversion of ATP into PPi after blood collection.

**Figure 3.**
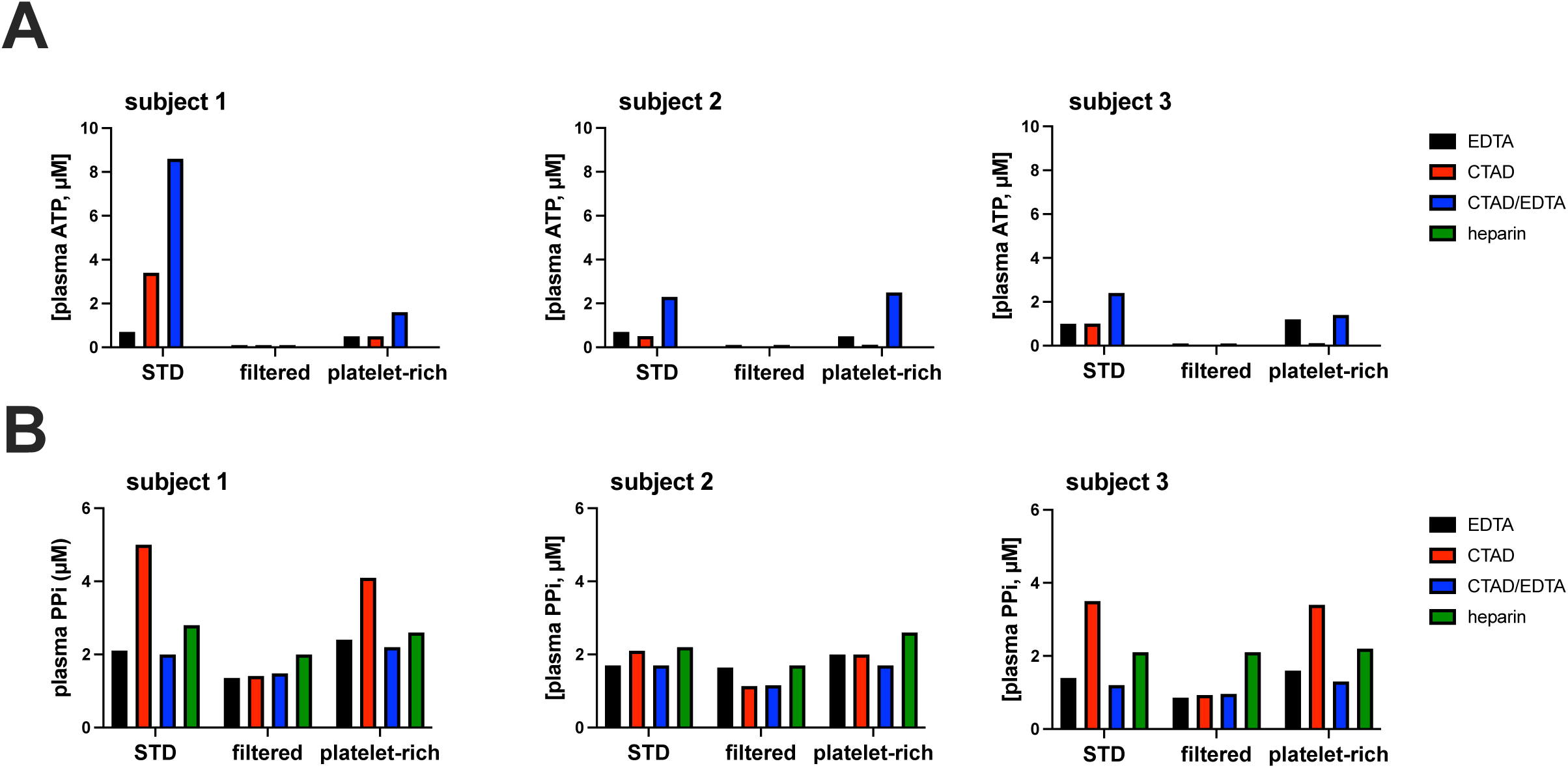
Blood processing affects the concentrations of ATP and PPi detected in plasma. Blood was collected from 3 participants via puncture of the antecubital vein and uncoagulated using the indicated anticoagulants. After preparation of plasma, ATP (A) and PPi (B) were determined in plasma not further processed (“STD”), in plasma that was filtered over a 300,000 mwco filter (“filtered”) and in platelet-rich plasma (“platelet-rich”). CTAD: citrate, theophylline, adenosine dipyridamole. EDTA: ethylenediaminetetraacetic acid.

The assay showed excellent reproducibility and interday variability in filtered EDTA, CTAD and CTAD/EDTA plasma samples of participant 3 (**Fig. 4**). ATP was only stable when EDTA was present, with CTAD plasma being completely free of ATP one day after storage at room temperature (**Fig. 4A**). The lower ATP concentrations detected in CTAD/EDTA plasma suggest some ATP degradation before EDTA addition. Notably, the EDTA-only plasma was prepared from blood collected in EDTA vacuum tubes, assuring rapid inactivation of ectonucleotidases. Following PPi concentrations in filtered heparin plasma from the same participant during storage at 22 °C, we first detected a substantial decline, then a clear increase, and finally a steady decline (**Fig. 4A**). Similar results, though less pronounced, were obtained with filtered heparinized plasma samples of participant 2 (**Supplemental figure 1**) stored at room temperature. We will come back to these unexpected results in the discussion section.

**Figure 4.**
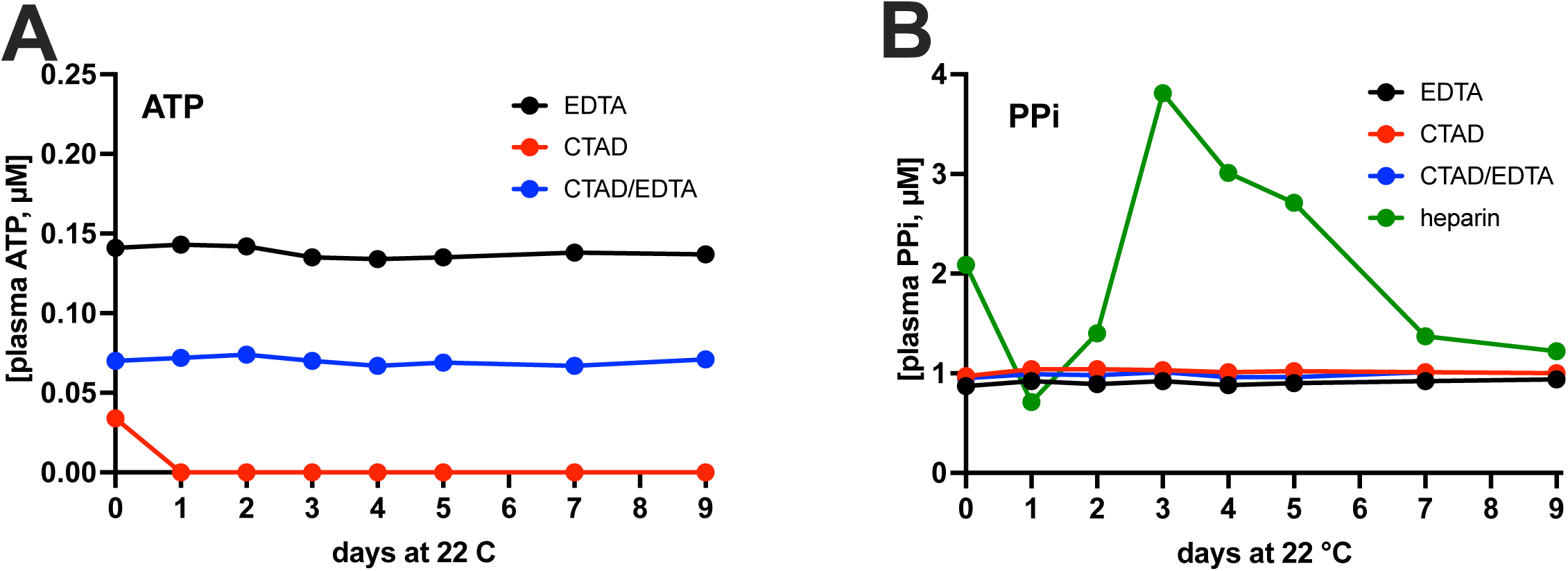
Stability of PPi and ATP in plasma depends on the used anticoagulant. Filtered plasma of participant 3, prepared using the indicated anticoagulants, was stored at room temperature (22 °C) and ATP (**A**) and PPi (**B**) were quantified at the indicated days. Of note, ATP was below the limit of detection in heparin plasma (see Figure 3B).

In PXE patients and *Abcc6*^-/-^ mice, absence of ABCC6 in the liver results in a 60-70% reduction of plasma PPi^5,6^. This reduction in plasma PPi is caused by the absence of ABCC6-mediated ATP release from the liver^6,29^. We next studied how well the optimized assay was able to detect the ABCC6-dependent difference in plasma PPi concentration in rats, a relatively new rodent model for PXE^26^. CTAD/EDTA was used as an anticoagulant in these experiments. The optimized assay returned similar average plasma PPi concentrations but yielded substantially lower variability within experimental groups resulting in a smaller overall p-value than the assay we used for our previous work^6,26^ (**Fig. 5**). The total blood volume of rats allowed to fill multiple CTAD tubes with blood from the same animal. The PPi concentrations in these sequentially drawn blood samples are given in Table III. There was considerable variability in these sequentially drawn samples in some animals. When blood was collected from the rats, the experiments shown in **Figure 4** had not yet been done and we were therefore unaware of the unexpected fact that citrate does not completely block conversion of ATP into PPi in whole blood. The variability in PPi concentration in sequentially drawn blood samples from the same animal is most likely caused by differences in the time between blood collection in the CTAD tubes and the subsequent addition of EDTA: In the absence of EDTA, extracellular ATP continues to be converted into PPi and, hence, results in increased plasma values.

**Table III:**
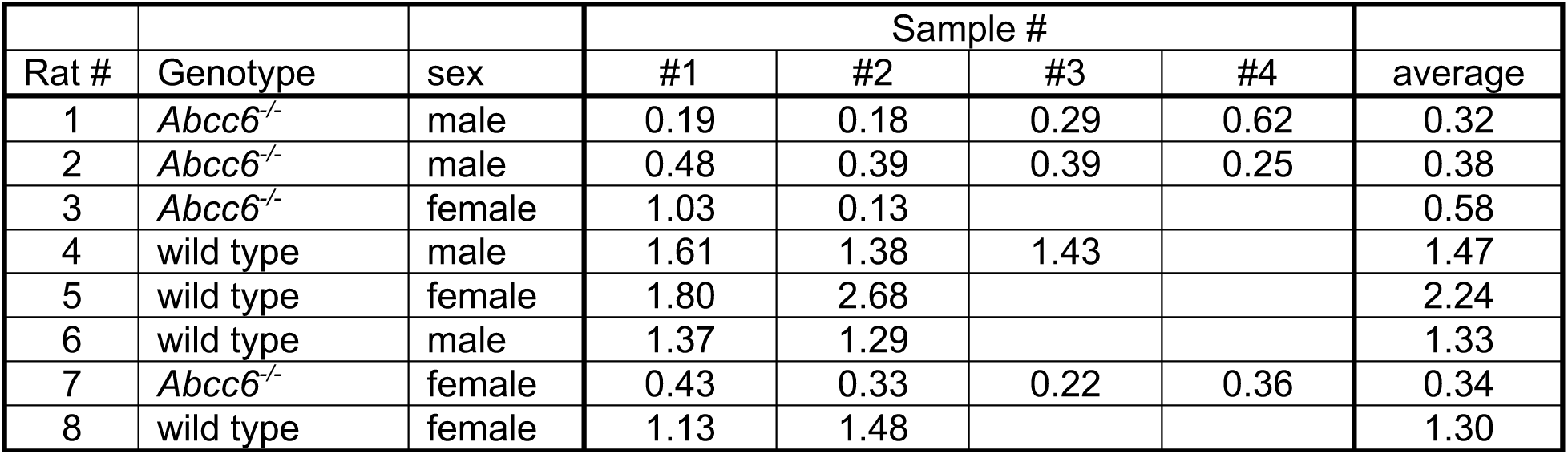
Sequentially taken blood samples collected from the same animal variation in plasma PPi concentrations. Blood was collected from *Abcc6*^-/-^ and wild type (Sprague Dawley) rats by cardiac puncture in CTAD tubes. Sequential blood samples were taken from the same animal till blood stopped flowing. After blood collection, 50 µl of a 15% K_3_EDTA solution was added per CTAD tube and plasma was prepared by centrifugation and filtration over a 300,000 Da mwco membrane.

| Rat # | Genotype | sex | Sample # |  |  |  | average |
| --- | --- | --- | --- | --- | --- | --- | --- |
|  |  |  | #1 | #2 | #3 | #4 |  |
| 1 | *Abcc6*<sup>-/-</sup> | male | 0.19 | 0.18 | 0.29 | 0.62 | 0.32 |
| 2 | *Abcc6*<sup>-/-</sup> | male | 0.48 | 0.39 | 0.39 | 0.25 | 0.38 |
| 3 | *Abcc6*<sup>-/-</sup> | female | 1.03 | 0.13 |  |  | 0.58 |
| 4 | wild type | male | 1.61 | 1.38 | 1.43 |  | 1.47 |
| 5 | wild type | female | 1.80 | 2.68 |  |  | 2.24 |
| 6 | wild type | male | 1.37 | 1.29 |  |  | 1.33 |
| 7 | *Abcc6*<sup>-/-</sup> | female | 0.43 | 0.33 | 0.22 | 0.36 | 0.34 |
| 8 | wild type | female | 1.13 | 1.48 |  |  | 1.30 |

**Figure 5.**
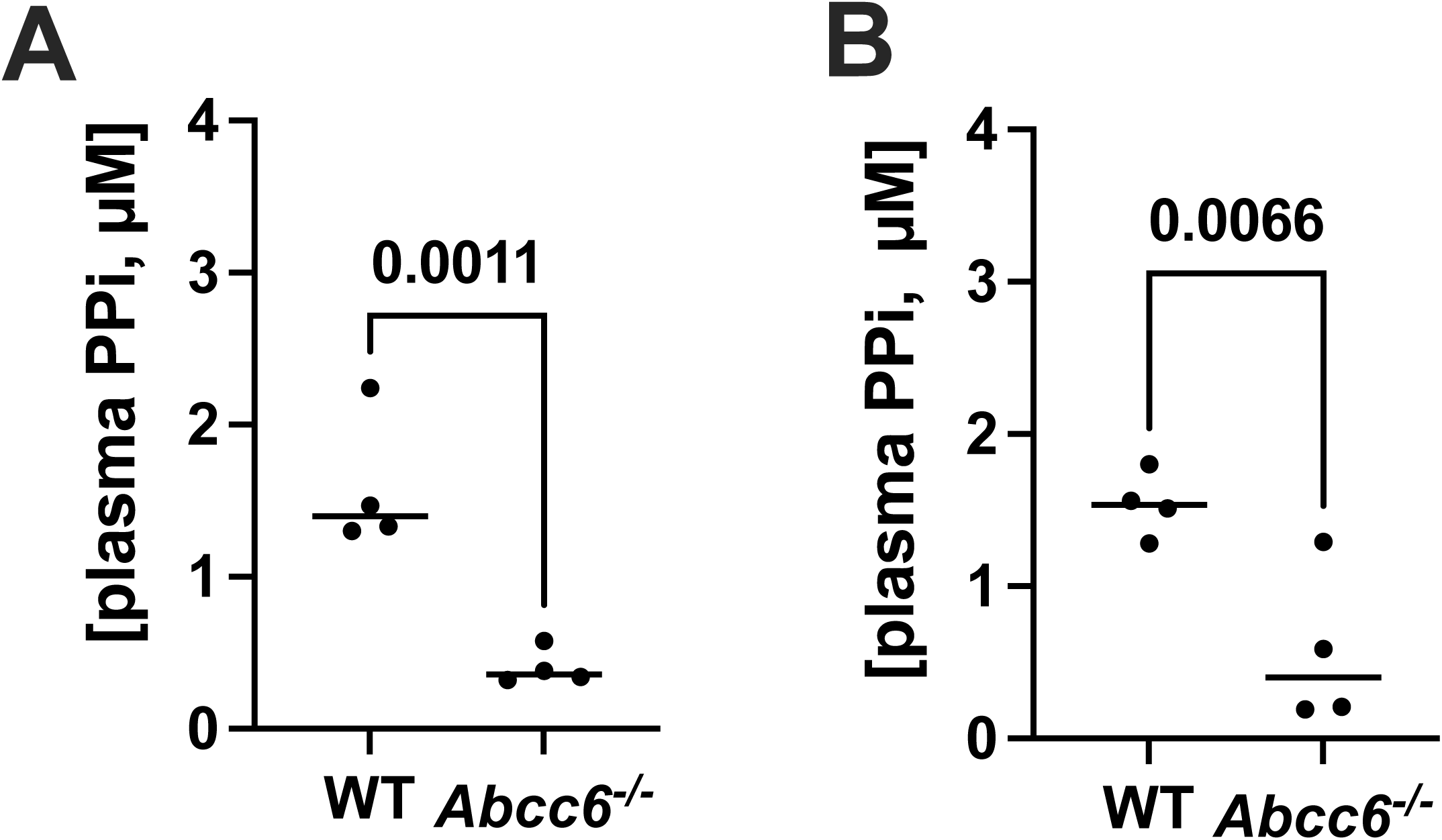
Comparison of the performance of the new (A) and old (B) ATPS-based PPi assay on plasma samples collected from wild type (WT) and *Abcc6*^-/-^ rats. Blood was collected from rats under terminal isoflurane anaesthesia by cardiac puncture in CTAD vacuum tubes. Directly after collection 50 µL of a 15% EDTA solution was added, and filtered plasma was prepared as detailed in the materials and methods section. PPi concentrations were determined in the resulting filtered plasma using the old and new assay. Each group contained 2 male and 2 female animals. Data represent individual data points and mean.

## Discussion

PPi plays a crucial role in the prevention of soft connective tissue mineralization^1^. Moreover, it is abundantly present in the mineral phase of bone^7,8,24^, where its exact function is still elusive. Several studies have recently explored approaches to increase circulating levels of PPi to counteract ectopic calcification^18,30–32^. A significant obstacle in furthering these studies is the large variability in different laboratories’ PPi concentrations detected in normal plasma. Moreover, assays relying on radionuclides are cumbersome and expensive. Therefore, we set out to develop an assay that did not employ radionuclides, which could still give reproducible and robust results. ATPS converts PPi in a one-step enzymatic reaction into ATP^33^. Luciferase allows detection of minute amounts of ATP but is prone to inhibition^34^. Interference by luciferase inhibitors was addressed in the current assay by using a more sensitive tube luminometer to quantify the luminescent signal which allowed dilution of plasma and by employing an internal ATP standard. Tube luminometers are about two orders of magnitude more sensitive than plate luminometers, meaning more diluted samples can be used, and issues related to luciferase inhibition can be reduced. The internal ATP standard corrects for any remaining inhibition. Notably, inhibition is not an issue for conversion of PPi into ATP by ATPS, as it would only delay the time until all PPi is converted.

The ATPS-based assay was highly sensitive, with a limit of detection (LOD) of at least 9 nM. Moreover, the assay took less than 5 minutes per plasma sample and showed good reproducibility and excellent recovery of spiked ATP/PPi. Collectively, these data indicate that the assay provides highly accurate and robust results as confirmed in human and rat plasma samples. We envision that if widely implemented in PPi plasma analysis, this assay would allow for the direct comparison of results obtained in different laboratories. The analyses of rat plasma showed that compared with the ATPS-based assay we used in previous work, our current assay showed reduced variability within experimental groups, suggesting higher accuracy. In future studies, this lower variability should allow for a reduction in sample size, e.g., fewer animals or patients per experimental group, while still retaining enough power to detect statistically significant differences. The high sensitivity of the used tube luminometer, which was used, allows for detecting PPi in complex matrices, like highly acidic bone extracts, even when the samples are quite dilute^7,8^. We expect the assay will perform equally well on other complex PPi-containing matrices, like teeth^35^. As the assay relies on the conversion of PPi into ATP, in its current form it performs less well on samples containing high amounts of ATP. This will, for instance, prevent determining intracellular PPi concentrations. A potential solution would be to degrade ATP using apyrase. Future work should reveal if it is possible to specifically deplete samples from ATP without affecting PPi concentrations.

Analyses of rat and human plasma demonstrate that the type of assay used is not the only factor contributing to variability in PPi concentrations detected in plasma. The technique used for blood collection, the anticoagulant used, and the processing of prepared plasma all affected PPi plasma concentrations. Using an anticoagulant that strongly chelates Mg^2+^ and Ca^2+^ seems crucial, as this quickly blocks conversion of any ATP released by cells into the extracellular environment (i.e. into plasma) after blood collection. Most laboratories use currently use CTAD as an anticoagulant when PPi needs to be quantified in plasma^17,18,36–39^. However, our data show that EDTA is the anticoagulant of choice, with citrate clearly allowing PPi formation after blood collection. This notion is supported by the striking stability of ATP and PPi in EDTA plasma stored at room temperature. This also demonstrates that ATP and PPi degradation requires divalent cations and is almost entirely enzymatic in nature. Our results indicate that heparin did not block ATP degradation and conversion into PPi, as apparent from the complete absence of ATP and the higher PPi concentrations detected in heparinized plasma samples. Stored at room temperature, PPi concentration detected in heparinized plasma showed an unexpected pattern over time. First, a decrease was seen, possibly due to the known TNAP activity in plasma^9^. The subsequent increase in plasma PPi concentrations after prolonged storage at room temperature is more difficult to explain. Filtered plasma was used in these experiments (mwco 300,000). Therefore, ATP release from a membrane-enclosed compartment with subsequent conversion into PPi cannot underlie these increased concentrations. Potential explanations include ENPP1-mediated conversion of polyphosphate, known to be present in plasma^40^, into PPi and slow release of protein-bound PPi^41^.

The multiple CTAD/EDTA blood tubes collected from individual rats showed substantial variability in plasma PPi concentrations in some animals. The samples were collected by inserting a dedicated blood collection needle into the heart and sequentially filling CTAD vacuum tubes without removing the needle. Differences in the time between blood sampling and addition of EDTA, differences in time before the blood samples were put on ice, or other slight alterations in the blood collection procedure might have contributed to the found variability. These data clearly indicate that blood collection and processing technique affects plasma PPi concentration. We expect that the points mentioned above also contribute to variability in plasma PPi concentrations detected in humans. Implementation of a detailed standardized procedure, therefore, seems critical to reducing variability.

ATP was virtually absent from filtered plasma, irrespective of the anticoagulant used. This indicates that in plasma ATP is present in a membrane-enclosed compartment or bound to a high molecular weight plasma protein. Lower levels of PPi were detected after filtration of plasma. Platelet dense granules contain high concentrations of ATP and PPi^28^. An attractive explanation is that the absence of ATP and reduced PPi concentrations in filtered plasma result from platelet removal. These data align well with previous studies exploring the effect of filtration on plasma PPi concentrations^12,42^ and suggest that using filtered plasma will reduce variability and provides a better estimate of the free PPi concentration in plasma.

In conclusion, we here describe a rapid, precise, accurate and reproducible assay to determine PPi in biological matrices. Analyses of rat and human plasma samples indicate that blood collection, anticoagulant and filtration all affect the amount of PPi measured. To compare plasma PPi concentrations quantified in different laboratories, it is crucial to standardize blood collection and processing, possibly using the data presented in this study as a guideline. This seems especially important to determine normal plasma PPi concentrations in adult and pediatric individuals.

## Data Availability

All data produced in the present study are available upon reasonable request to the authors

## Acknowledgments

This research was funded by National Institutes of Health, Grant R01AR072695 (K.v.d.W.), U.S. Department of State (Fulbright Visiting Scholar Program), National Research, Development and Innovation Office (OTKA FK131946), Hungarian Academy of Sciences (Bolyai János Fellowship BO/00730/19/8, Mobility grant) and the Ministry for Innovation and Technology from the source of the National Research, Development and Innovation Fund (ÚNKP-2021 New National Excellence Program) to F.S. Further funding for this work was provided by PXE International for K.v.d.W. and F.S. We are grateful to individuals affected by pseudoxanthoma elasticum (PXE) and their continued support of our research.

## Legends

**Figure S1. Stability of PPi in heparinized plasma of subject 2**. Filtered, heparinized plasma of subject 2, was stored at room temperature (22 °C) and PPi was quantified at the indicated days. Of note, ATP was below the limit of detection in heparin plasma (see Figure 3B).

