## Supplementary figures and images for "A new enzymatic assay to quantify inorganic pyrophosphate in plasma"

### Supplemental Figure 1

#### Supplemental Figure 1

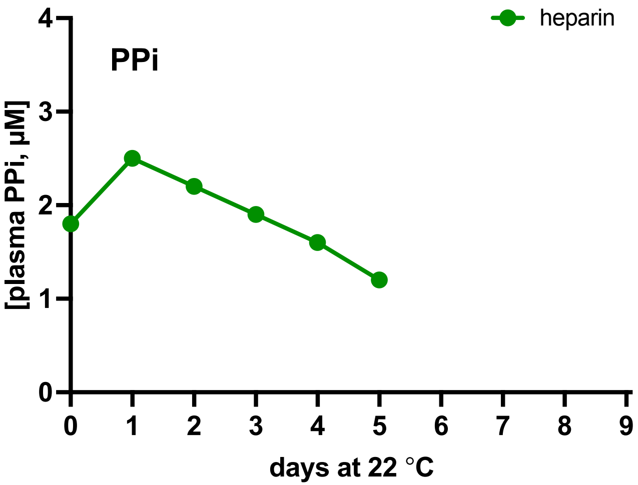
